# BrainAgeNeXt: Advancing Brain Age Modeling for Individuals with Multiple Sclerosis

**DOI:** 10.1101/2024.08.10.24311686

**Authors:** Francesco La Rosa, Jonadab Dos Santos Silva, Emma Dereskewicz, Azzurra Invernizzi, Noa Cahan, Julia Galasso, Nadia Garcia, Robin Graney, Sarah Levy, Gaurav Verma, Priti Balchandani, Daniel S Reich, Megan Horton, Hayit Greenspan, James Sumowski, Merixtell Bach Cuadra, Erin S Beck

**Affiliations:** Department of Neurology, Icahn School of Medicine at Mount Sinai, New York, NY, 10029, USA; Department of Environmental Medicine and Public Health, Icahn School of Medicine at Mount Sinai, New York, NY, 10029, USA; BioMedical Engineering and Imaging Institute, Icahn School of Medicine at Mount Sinai, New York, NY, USA; Translational Neuroradiology Section, National Institute of Neurological Disorders and Stroke, National Institutes of Health, Bethesda, MD, USA; CIBM Center for Biomedical Imaging, Switzerland; Radiology Department, University of Lausanne and Lausanne University Hospital, Switzerland

## Abstract

Aging is associated with structural brain changes, cognitive decline, and neurodegenerative diseases. Brain age, an imaging biomarker sensitive to deviations from healthy aging, offers insights into structural aging variations and is a potential prognostic biomarker in neurodegenerative conditions. This study introduces BrainAgeNeXt, a novel convolutional neural network inspired by the MedNeXt framework, designed to predict brain age from T1-weighted magnetic resonance imaging (MRI) scans. BrainAgeNeXt was trained and validated on 11,574 MRI scans from 33 private and publicly available datasets of healthy volunteers, aged 5 to 95 years, imaged with 3T and 7T MRI. Performance was compared against three state-of-the-art brain age prediction methods. BrainAgeNeXt achieved a mean absolute error (MAE) of 2.78 ± 3.64 years, lower than the compared methods (MAE = 3.55, 3.59, and 4.16 years, respectively). We tested all methods also across different levels of image quality, and BrainAgeNeXt performed well even with motion artifacts and less common 7T MRI data. In three longitudinal multiple sclerosis (MS) cohorts (273 individuals), brain age was, on average, 4.21 ± 6.51 years greater than chronological age. Longitudinal analysis indicated that brain age increased by 1.15 years per chronological year in individuals with MS (95% CI = [1.05, 1.26]). Moreover, in early MS, individuals with worsening disability had a higher annual increase in brain age compared to those with stable clinical assessments (1.24 vs. 0.75, p < 0.01). These findings suggest that brain age is a promising prognostic biomarker for MS progression and potentially a valuable endpoint for clinical trials.

## Introduction

The aging population presents a significant global challenge, particularly concerning brain health^1^. As individuals age, the brain undergoes profound structural and functional transformations. These include reductions in brain volume, notably in the prefrontal cortex and hippocampus, and the accumulation of white matter lesions, all of which contribute to a decline in memory, executive function, and overall cognitive capabilities^2^. The incidence of neurodegenerative diseases, such as Alzheimer’s disease and Parkinson’s disease, increases with age, further contributing to cognitive deterioration. As the aging population expands, it becomes crucial to deepen our understanding of these neurobiological changes in order to develop effective interventions that can preserve cognitive function and improve the quality of life for the elderly and those affected by neurodegenerative conditions.

Brain age (BA) is a biomarker estimated from magnetic resonance imaging (MRI) that detects deviations from healthy aging^3^. The gap between chronological age (CA) and estimated BA is known as brain age difference (BAD). Individuals with a large, positive BAD might be undergoing accelerated aging, while those with a negative BAD might have a healthier brain relative to their chronological age. Brain age can also serve as an imaging biomarker in neurologic and psychiatric diseases. Studies have shown that BAD tends to be higher in people with psychiatric and neurological conditions, including Alzheimer’s disease, multiple sclerosis (MS), and Parkinson’s disease, compared to healthy volunteers (HV)^4,5^.

MS is an autoimmune disease characterized by inflammatory demyelination and progressive neurodegeneration, leading to physical and cognitive impairment^6^. Chronological age is strongly associated with the clinical course of the disease^7^. Individuals who are younger at clinical disease onset are more likely to have a predominantly inflammatory disease course with more total relapses before entering the progressive phase of the disease^8^. This transition from a relapsing-remitting to a progressive clinical course in MS occurs for some but not all individuals. Despite a growing understanding of the disease course and pathophysiological mechanisms, the currently available clinical and biomedical tools are still limited in their ability to identify those at higher risk for progressive disease. With aging, remyelination is less effective, and the restorative efficacy from MS relapses declines alongside reduced plasticity^9,10^. Older individuals with MS also experience increased rates of brain atrophy^11^, which are moderately associated with physical disability and cognitive impairment^12^. Currently, MS clinical care relies heavily on MRI findings^13^. However, MRI biomarkers, including lesion burden and brain atrophy, have limited utility in predicting cognitive decline and disease progression independent of relapse activity (PIRA). PIRA, often seen in people with MS (pwMS), is likely driven in part by neurodegeneration^14^. There is currently a lack of effective outcome measures for proof-of-concept clinical trials in progressive MS, where disability accumulation is too slow to be a feasible assessment metric^15^.

Most BA prediction models applied to MS have traditionally relied on classical machine learning methods, which are trained using previously extracted brain volumetric features^5,16,17^. This approach typically involves segmenting the brain into various structures and then extracting volumetric or morphological features from these regions using brain imaging software such as FreeSurfer^18^ or FSL^19^. However, brain parcellation tools often yield inconsistent results when applied to MRI scans obtained from different sites or using different imaging parameters^20,21^. Thus, relying on pre-extracted brain volumetric features can limit the applicability of these models to real-world clinical settings, where scans from various sources and conditions are common, or to research studies that include multi-site data. Additionally, while volumetric features provide useful information, they may only partially represent the intricate patterns of brain aging across the human lifespan.

In recent years, convolutional neural networks (CNNs) have demonstrated exceptional performance in image analysis tasks, including medical imaging. These networks can automatically learn and extract relevant features from high-dimensional data, making them competitive for complex tasks such as brain age prediction. CNNs provide a more flexible and robust alternative by directly learning from raw and heterogenous MRI data without relying on several preprocessing steps or predefined features^22–25^. Recent state-of-the-art brain age models have employed advanced deep learning techniques, achieving high accuracy and robustness in their predictions^26^. These models leverage large datasets and architectures like CNNs and graph neural networks to capture detailed patterns in MRI data^23,27^, significantly outperforming traditional machine learning methods^28^.

In this study, we propose a novel CNN for the BA estimation task. By utilizing the advanced capabilities of CNNs, we aim to overcome the limitations associated with classical ML approaches and improve the robustness and accuracy of brain age estimation in MS. Brain age estimation has mostly been explored at 1.5T and 3T^28,29^. While 7T MRI has led to important insights into neurologic diseases, including MS^30,31^, only one study to date has explored the BA measurement using 7T MRI^32^. Verma et al. proposed a regression-based model for predicting BA from 7T MRI volumetric features quantified using FreeSurfer^18^. In our study, we include 7T MRI data in the training and testing datasets of our deep learning-based model, leveraging its enhanced resolution and tissue contrast to potentially improve the BA prediction accuracy.

In this work, we analyze a substantial dataset of over 11,000 1.5T, 3T, and 7T MRI scans, spanning most of the human lifespan, to characterize typical aging patterns within the healthy population. Our contribution is twofold: first, we propose BrainAgeNeXt, a novel CNN architecture inspired by the MedNeXt framework, for estimating brain age from diverse T1-weighted (T1w) MRI scans. We evaluate BrainAgeNeXt on large, multi-site cohorts of HV imaged with both 3T and 7T MRI. Second, we extend the application of the brain age paradigm to three longitudinal cohorts of subjects with MS to characterize brain age in MS.

## Datasets

### Healthy subjects

We aggregated several publicly available and private datasets of HV imaged at 1.5T, 3T, and 7T, to obtain a dataset of 11574 T1w MRI (Figure 1). The HV MRI scans used for training and validation (*n* = 10,051, from 8,838 unique subjects) are shown in Table 1. The age range is 5-95 years (mean 39.2 ± 24.0 years), and 51% are females. The training set included MRI images acquired at 75 sites with various scanners and imaging protocols. Various T1w MRI sequences were included: spoiled gradient recalled echo (SPGR), magnetization-prepared rapid gradient echo (MPRAGE), and magnetization-prepared 2 rapid acquisition gradient echoes (MP2RAGE)^33^. For testing, we selected 1,523 MRI scans from 10 sites, unseen during training, with 3T or 7T MRI acquired with Siemens, Philips, or GE scanners. Some datasets contained two scans of a single subject, either at different time points (n = 50) or magnetic field strengths (n = 9).. The age range is 8-89 years (mean 39.1 ± 20.9 years), and 59% are females. Table 2 summarizes the main characteristics of the testing dataset. A histogram of the age distribution for both the training and testing datasets is shown in Figure 2B.

**Figure 1:**
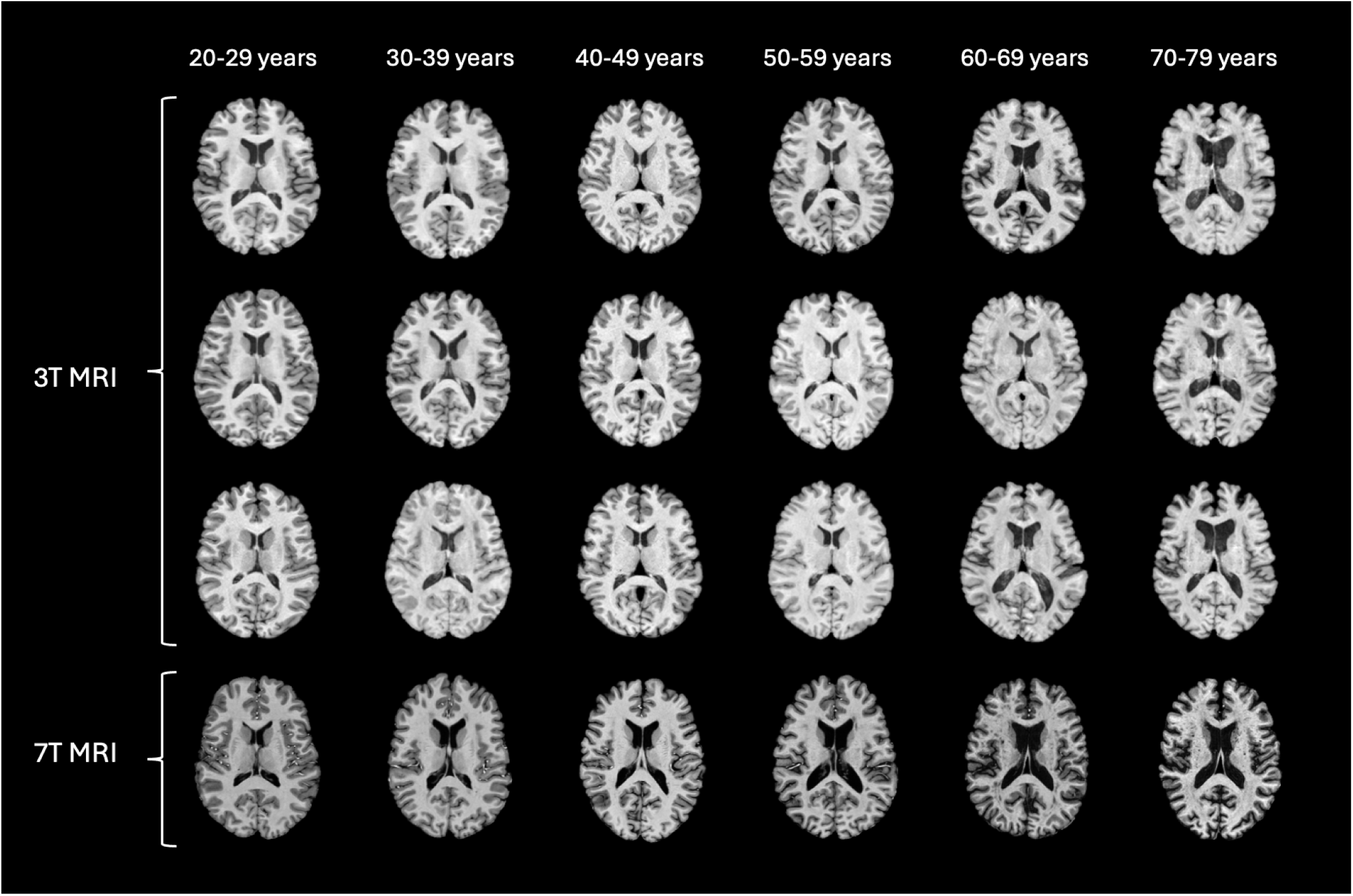
Representative axial MRI slices after preprocessing from healthy subjects across different decades. The image illustrates age-related anatomical changes. Different T1w sequences are shown for the 3T images, whereas the 7T scans are all MP2RAGE.

**Figure 2.**
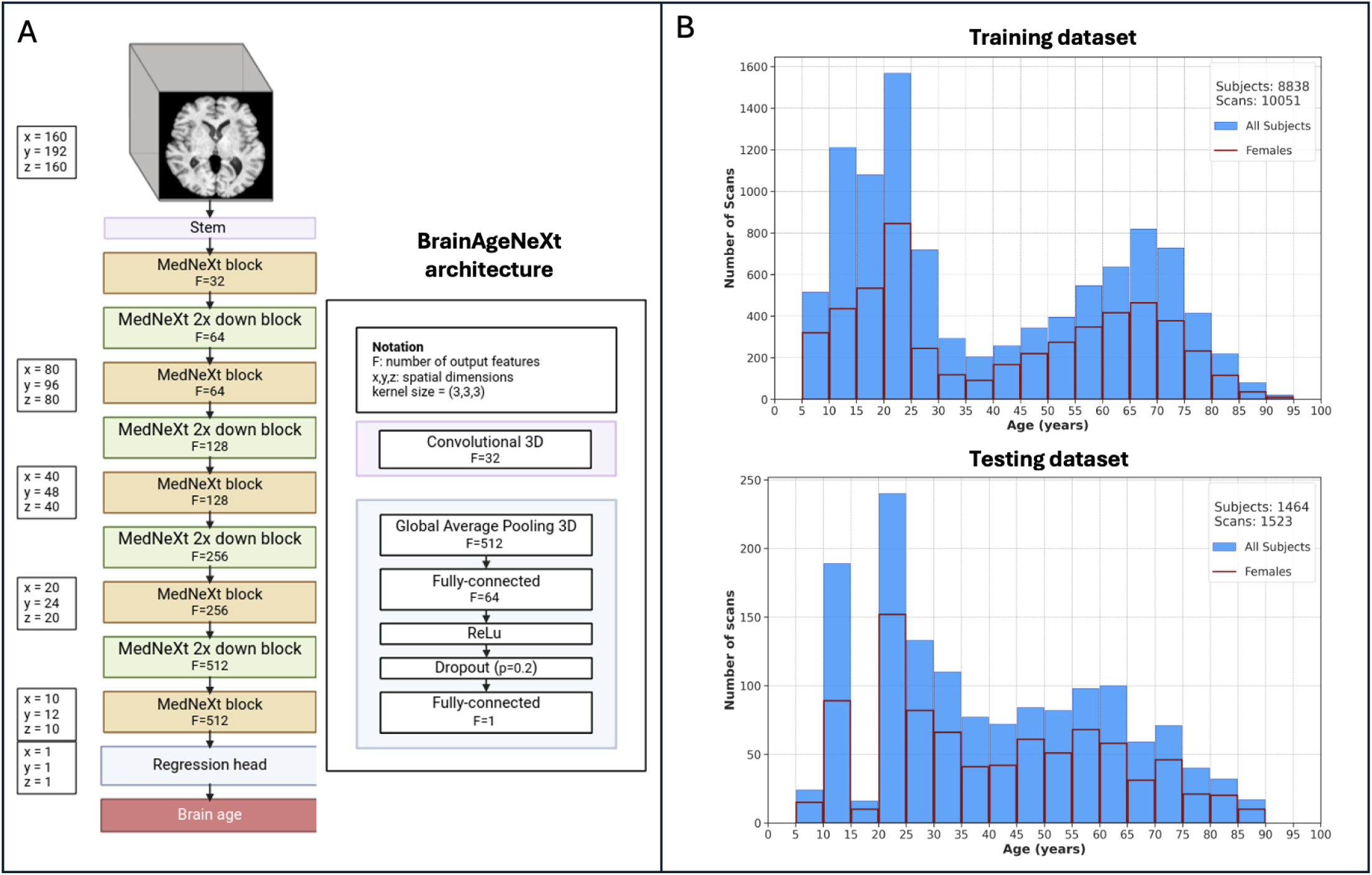
BrainAgeNeXt architecture and training/testing datasets. A: Scheme of the proposed BrainAgeNeXt architecture. B: Histograms showing the age distribution of the HV training and testing MRI scans.

**Table 1:** Datasets used for training BrainAgeNeXt. MPRAGE: Magnetization Prepared Rapid Gradient Echo. MP2RAGE: Magnetization Prepared 2 Rapid Gradient Echoes.

| Training Datasets | Field Strength | T1w Sequence | Number of subjects (age range) | Sex (F) |
| --- | --- | --- | --- | --- |
| openBHB<sup>29</sup> | 1.5T, 3T | T1w, MPRAGE | 5,330 (5-86y) | 54% |
| Cam-CAN<sup>36</sup> | 3T | MPRAGE | 653 (18-87y) | 49% |
| Oasis 3<sup>37</sup> | 3T | MPRAGE | 755 (42-95y) | 48% |
| Oasis 4<sup>38</sup> | 3T | MPRAGE | 663 (21-94y) | 54% |
| QTAB<sup>39</sup> | 3T | MPRAGE | 510 (9-16y) | 52% |
| NKI-RS<sup>40</sup> | 3T | MPRAGE | 653 (6-85y) | 51% |
| PREVENT-AD<sup>41</sup> | 3T | MP2RAGE | 43 (57-82y) | 51% |
| ATAG<sup>42</sup> | 7T | MP2RAGE | 53 (19-75y) | 64% |
| CEREBRUM-7T<sup>43</sup> | 7T | MP2RAGE | 76 (19-40y) | 48% |
| CoRR<sup>44</sup> | 7T | MP2RAGE | 22 (22-30y) | 45% |
| CFFM<sup>45</sup> | 7T | MP2RAGE | 32 (20-66y) | 47% |
| Mount Sinai<sup>46</sup> | 7T | MP2RAGE | 48 (20-70y) | 55% |

**Table 2:** Testing datasets. ^1^3T images were acquired at a 1.0-1.5mm resolution, 7T images had a 0.5-0.8mm resolution. MPRAGE: Magnetization Prepared Rapid Gradient Echo. MP2RAGE: Magnetization Prepared 2 Rapid Gradient Echoes. SPGR: Spoiled Gradient Recalled Echo.

| Dataset | Field Strength | Scanner | T1w Sequence<sup>1</sup> | Number of subjects (age range) | Sex (% female) |
| --- | --- | --- | --- | --- | --- |
| MR-ART<sup>47</sup> | 3T | Siemens Prisma | MPRAGE | 148 (18–75y) | 64% |
| DLBS<sup>48</sup> | 3T | Philips | MPRAGE | 315 (10-81y) | 63% |
| SALD<sup>49</sup> | 3T | Siemens Tim Trio | MPRAGE | 493 (19-80y) | 62% |
| Horta et al.<sup>50</sup> | 3T | Philips Achieve | MPRAGE | 87 (18-81y) | 51% |
| PROGRESS | 3T | Philips whole-body system | MPRAGE | 215 (8-15y) | 49% |
| RADIEMS<sup>34</sup> | 3T | Siemens Skyra | MPRAGE | 32 (24-49y) | 64% |
| NIMH<sup>51</sup> | 3T | GE Discovery MR750 | SPGR | 50 (21-67y) | 47% |
| NIMH<sup>51</sup> | 3T | GE Discovery MR750 | MPRAGE | 68 (21-71y) | 46% |
| Liu et al.<sup>52</sup> | 3T | Siemens Prisma | MPRAGE | 44 (19-45y) | 66% |
| Chen et al.<sup>53</sup> | 3T | Siemens Prisma | MPRAGE | 10 (25-41y) | 30% |
| Chen et al.<sup>53</sup> | 7T | Siemens Terra | MP2RAGE | 9 (25-41y) | 33% |
| CBS<sup>54</sup> | 7T | Siemens Magnetom | MP2RAGE | 28 (21-36y) | 54% |
| NIH<sup>55</sup> | 7T | Siemens Magnetom | MP2RAGE | 5 (40-78y) | 60% |
| Mount Sinai | 7T | Siemens Magnetom | MP2RAGE | 19 (25-76y) | 55% |

### Subjects with multiple sclerosis

Three longitudinal cohorts of pwMS were included in this study:

● Reserve Against Disability in Early Multiple Sclerosis (RADIEMS, Mount Sinai)^34^: a longitudinal study investigating factors associated with disability progression in pwMS diagnosed within 5 years at the time of enrollment. 185 pwMS (165 relapsing remitting MS (RRMS), 20 clinically isolated syndrome; disease duration 2.2 ± 1.4 years; mean age at enrollment 34 years (range 20-50); 67% female) underwent 3T MRI and clinical assessment, which included extensive cognitive testing at baseline and at year 3 follow-up visits.
● Longitudinal Cortical Lesion study (LCL, National Institutes of Health [NIH])^35^: 64 pwMS (45 RRMS, 19 progressive MS; disease duration 14.2 ± 10.9 years; mean age 46 years (range 20–77); 64% female) underwent 3T and 7T MRI and clinical assessment at baseline, year 1 and year 2. At baseline, the median interval between 3 and 7 T MRI was 9 weeks (range 1–34).
● Longitudinal 7T (Long 7T, Mount Sinai): longitudinal study of advanced lesional biomarkers in newly diagnosed pwMS with 7T MRI and clinical assessments. This includes 28 people with RRMS (disease duration 0.8 ± 0.4 years, mean age 34 years (range 24–54), 62% female).

For all cohorts, the clinical assessment included Expanded Disability Status Scale (EDSS), Symbol Digit Modalities Test (SDMT), Nine-Hole Peg Test (9HPT), and Timed 25-Foot Walk (T25FW). For the LCL cohort, the written version of the SDMT was performed, whereas the oral version was used for the RADIEMS and Long 7T cohorts.

## Methods

### Preprocessing

All T1w MRI scans underwent skull-stripping with SynthStrip^56^, N4 bias field correction with default parameters, and linear registration with 6 degrees of freedom to the MNI152 1mm isotropic space with ANTs^57^. This minimal preprocessing ensures that the images are in the same space, enabling consistent analysis across subjects.

### Network architecture

We propose BrainAgeNeXt, a CNN inspired by the MedNeXt framework^58^, which predicts brain age from minimally preprocessed 3D T1w MRI. BrainAgeNeXt architecture is composed of four MedNeXt blocks followed by a regression header. The MedNeXt blocks are a 3D extension of the ConvNeXt networks^59^, which incorporate architectural insights derived from Vision Transformers and Swin Transformers^60^ into a convolutional framework. Each MedNeXt block has 3 layers, mirroring a transformer. Additional details regarding these layers can be found in the original MedNeXt paper^58^. Following the fourth MedNeXt block, the regression head first applies 3D pooling to average features spatially and reduce their dimensionality. Finally, two fully connected layers are added to provide a unique number, brain age, as output. These last two layers use a ReLU activation function, and the first one has a dropout rate of 0.2.

### Data augmentation

Several intensity and spatial transformations were applied to the input data during training to improve the generalizability of BrainAgeNeXt. The spatial transformations consisted of random rotations along the three axes (up to 0.1 radiant), affine transformations, zooming in and out (up to 5% of the image size), and foreground cropping and padding to the input dimensions (160,192,160). The intensity augmentation included random contrast adjustment, gamma transformations, simulated bias field, random noise, and random swapping patches within the image. Moreover, random motion and ghosting artifacts were simulated^61^. All transforms were applied with a random 0.2 probability at each training iteration. MONAI^62^ and TorchIO^61^ python libraries were used for these transformations.

### Configuration

The network was trained for 200 epochs using the L2 loss function, with a batch size set to 4. The code is a custom Python script based on the MONAI framework^62^. Training was performed on one NVIDIA A100 80GB GPU and took approximately 6 days. From the training set (Table 1), we extracted both an internal and an external validation dataset. The internal validation set consisted of 5% of the training set, randomly sampled but balanced to ensure representation across each 5-year interval throughout the lifespan. The external validation dataset included MRI scans from an unseen site during training (N=498), covering an age range of 6–84 years. We saved the best model based on the lowest combination of internal and external validation losses, ensuring robust performance across diverse datasets. Our code is made publicly available^1^.

### Brain age prediction

The output provided by the network is corrected for the known regression to the mean bias, which causes BA overestimation for younger subjects and underestimation for older ones^3^. To achieve this, we applied the correction proposed by Beheshti et al.^63^:

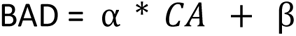

α and β are obtained by fitting a linear regression model into the validation dataset. The final BA prediction is obtained by taking the median of the outputs from five independently trained models with identical configurations but random weight initializations.

### Baselines

The DenseNet convolutional neural network architecture^64^ and its 3D extension have been extensively used for the BA estimation task in the literature^24,28^. We trained one DenseNet architecture as a baseline model to compare with our proposed BrainAgeNeXt using the same training dataset. BrainAgeNeXt, inspired by the ConvNeXt^59^ and MedNeXt^58^ architectures, differs from the baseline DenseNet by incorporating advanced convolutional techniques for improved feature extraction, performance, and generalization in brain age prediction. In addition, we tested two publicly available methods, one CNN and one regression-based model, that in a recent review resulted as the top performing BA estimation methods^28^. All three methods were tested on the same testing datasets as BrainAgeNeXt.

● **DenseNet**: Wood et al.^24,65^ proposed to train a DenseNet201 to estimate BA from routine clinical MRI data. When trained on over 19,000 MRI scans (18–96 years), the model achieved a mean age error (MAE) < 4.0 years, in line with other state-of-the-art approaches ^3,22,66^. We retrained this architecture using our training dataset (age 43.4 ± 19.3 years, range 5–95, 47% female) using the same data augmentation and validation strategy as for BrainAgeNeXt..
● **pyment**: Leonardsen et al. developed a shallow 3D convolutional neural network trained from skull-stripped T1w MRI registered to the MNI152 space^26^. FreeSurfer was used for skull-stripping and FSL *FLIRT* for linear registration. The network was trained with 53,542 HV from 21 datasets (age range 3–95 years, 52% female). For our evaluation, we applied the publicly available trained model provided.
● **brainageR**: A BA estimation method based on Gaussian process regression, a nonparametric Bayesian regression approach^67,68^. Raw T1w MRI scans were segmented with SPM12 into gray matter, white matter, and CSF. Their probability maps were normalized and concatenated. Following a principal component analysis, a Gaussian progress regression model was trained to predict BA. This method was trained on 2,277 HV (age 40.6 ± 21.4 years, range 18–92, 48% female). As some of these HV were included in our testing dataset, we evaluated brainageR only on the remaining scans (*n* = 352).

### Image processing

To determine the relationship between brain volumetric measures and BA predictions, tissue segmentation was performed on all HV testing images using SynthSeg (Freesurfer 7.4.1^18,56^). In RADIEMS, MS lesions were segmented automatically^69^ using the FLAIR images followed by manual correction. In LCL, MS lesions were segmented semiautomatically using FLAIR and T1w 3T images^35^. For all 3T scans of pwMS, lesion filling was performed using FSL^19^ within the white matter (WM) tissue mask from FreeSurfer. The lesion-filled images were then visually inspected, and if necessary, the WM masks were manually corrected, and the lesion-filling process was repeated. BrainAgeNeXt’s predictions from the original and lesion-filled images were then compared (see Results).

### Statistical analysis

To estimate the accuracy of BA prediction in HV, we computed the mean absolute error (MAE):

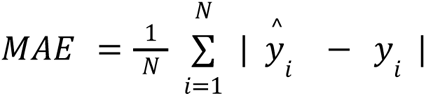

where N is the number of subjects, *ŷ_i_* is the BA prediction for subject i, and *y_i_* is their CA.

The MAE represents the average absolute difference between the predicted BA and the chronological age (CA). For healthy subjects, the MAE should be close to zero. Pearson’s *r* was calculated to assess the correlations between predicted BA and CA, as well as between predicted BA and BAD. Ideally, for an unbiased model, Pearson’s *r* for the correlation between CA and BAD (i.e., the difference between CA and BA) should be as close as possible to 0. Using the Mann-Whitney U test, we assessed whether the differences in the MAE between different brain age models are statistically significant, while also considering the effect size to understand the practical significance of these differences.

Monotonic correlations between MRI markers or clinical measures and BAD were evaluated using Spearman’s rank correlation. Statistical significance was determined using a *p*-value threshold of 0.05, adjusted for multiple comparisons using the false discovery rate (FDR) correction.

## Results

### Healthy volunteers

The results of the HV analysis are summarized in Table 3 and Figure 3. In the testing dataset, consisting of 1,523 HV, our proposed BrainAgeNeXt achieves the lowest MAE of 2.78 ± 3.64, followed by DenseNet (MAE = 3.55 ± 3.90 years), pyment (MAE = 3.59 ± 3.66 years), and brainageR (MAE = 4.46 ± 5.14 years). No statistically significant difference in BAD between males and females was observed for BrainAgeNeXt (*p* = 0.37). Analyzing 7T MRI scans separately (*n* = 61), the MAE for BrainAgeNeXt is 2.08, versus 4.12 for DenseNet (U = 1929, *p* = 0.73, compared to BrainAgeNeXt; with a rank-biserial correlation (r) = 0.52, indicating a large effect size) and 7.98 for pyment (U = 938, *p* < 0.001, compared to BrainAgeNeXt; r = 0.25, indicating a small effect size). Additionally, all models were compared on the MR-ART dataset, which presents three T1w scans for each of its 148 HV: one motionless, one with moderate motion artifacts, and one with stronger motion artifacts (Table 3).

**Figure 3.**
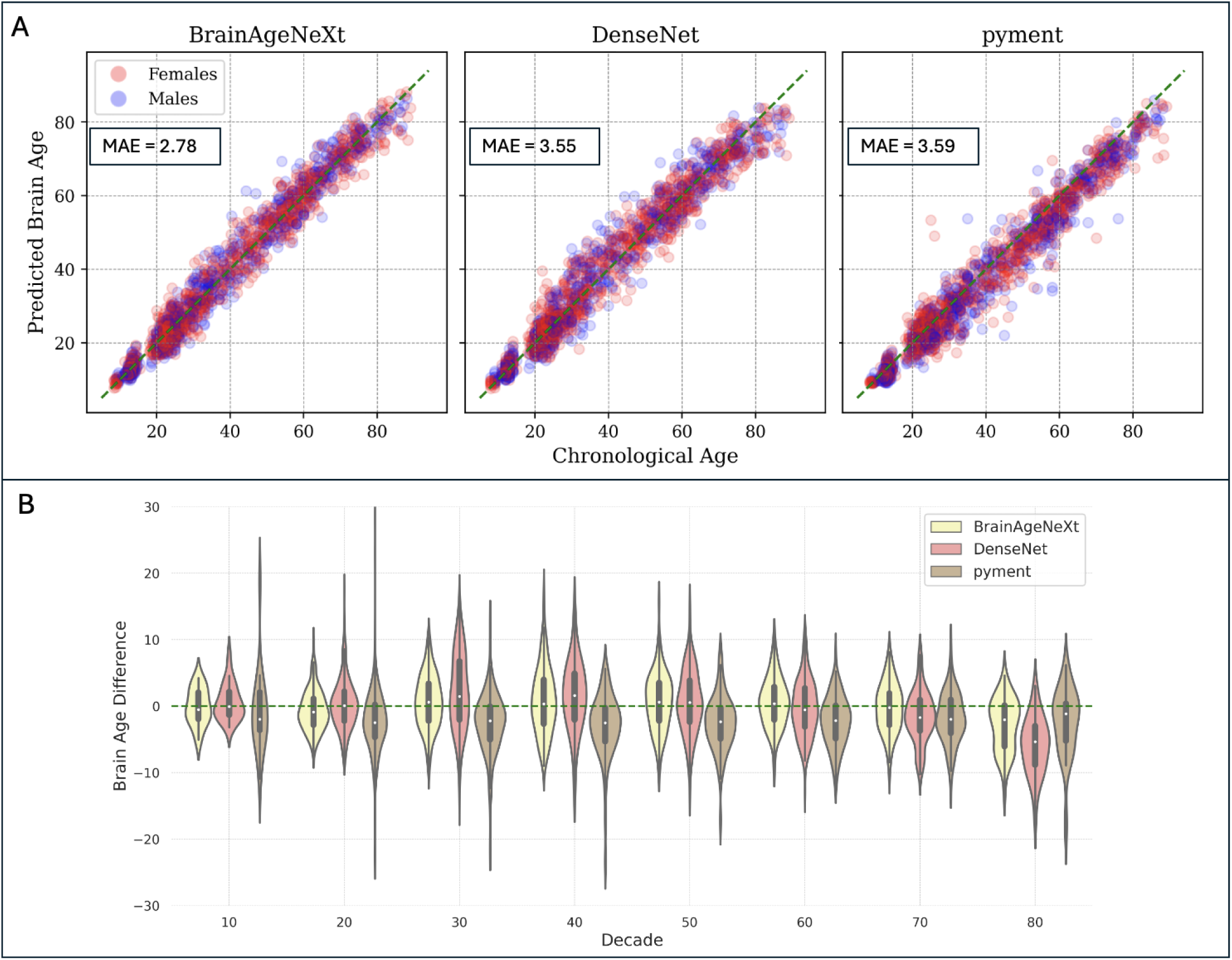
BrainAgeNeXt accurately predicts brain age in healthy volunteers. A: Scatter plots comparing predicted brain age versus chronological age for BrainAgeNeXt, DenseNet, and pyment in the testing dataset (N=1’523). B: Violin plot showing the BAD for the three methods across all decades.

**Table 3.** Performance of BrainAgeNeXt vs state-of-the-art models. MAE: mean age error. BA: brain age. CA: chronological age. BAD: brain age difference. Significance levels: * *p* < 0.05, ** *p* < 0.01, *** *p* < 0.001.

| | Testing dataset ($n = 1'523$) | | | 7T MRI ($n = 61$) | | |
| --- | --- | --- | --- | --- | --- | --- |
| Model | MAE (years) | BA vs CA<br>(Pearson's $r$) | BAD vs CA<br>(Pearson's $r$) | MAE (years) | BA vs CA<br>(Pearson's $r$) | BAD vs CA<br>(Pearson's $r$) |
| BrainAgeNeXt | **2.78** | **0.985\*\*\*** | **-0.010** | **2.08** | **0.982\*\*\*** | **0.001** |
| DenseNet | 3.55 | 0.978*** | -0.212 | 4.12 | 0.967*** | -0.337 |
| pyment | 3.59 | 0.972 | -0.309 | 7.98 | -0.683 | 0.655 |
| brainageR* | 4.46 | 0.965 | -0.202 | 12.22 | -0.318 | 0.736 |

|  | MR-ART (N=148)<br>Motionless |  |  | MR-ART (N=148)<br>Moderate motion |  |  | MR-ART (N=148)<br>Strong motion |  |  |
| --- | --- | --- | --- | --- | --- | --- | --- | --- | --- |
| Model | MAE<br>(years) | BA vs CA<br>(Pearson's $r$) | BAD vs CA<br>(Pearson's $r$) | MAE<br>(years) | BA vs CA<br>(Pearson's $r$) | BAD vs CA<br>(Pearson's $r$) | MAE<br>(years) | BA vs CA<br>(Pearson's $r$) | BAD vs CA<br>(Pearson's $r$) |
| BrainAgeNeXt | **2.82** | **0.968\*\*\*** | **-0.043** | **3.16** | **0.947\*\*\*** | **-0.022** | 3.50 | **0.942\*\*\*** | 0.062 |
| DenseNet | 3.26 | 0.952*** | 0.061 | 3.49 | 0.945 | 0.163 | 3.98 | 0.932*** | **0.055** |
| pyment | 3.18 | 0.947 | -0.382 | 3.39 | 0.939 | -0.155 | 3.92 | 0.945 | -0.187 |
| brainageR | 3.17 | 0.970 | -0.230 | 5.48 | 0.830 | -0.325 | 7.41 | 0.811 | -0.336 |

To better understand BrainAgeNeXt predictions, we investigated the correlations between brain parcel volumes as obtained with FreeSurfer^56^ and BAD in the testing dataset (Figure 4). We combined parcels from the left and right hemispheres, and each parcel volume was normalized by the total intracranial volume. The strongest correlations with BAD were seen for the cerebral cortex (*r* = −0.23, *p* < 0.001), putamen (*r* = −0.21, *p* < 0.001), and cerebrospinal fluid (*r* = 0.20, *p* < 0.001).

**Figure 4.**
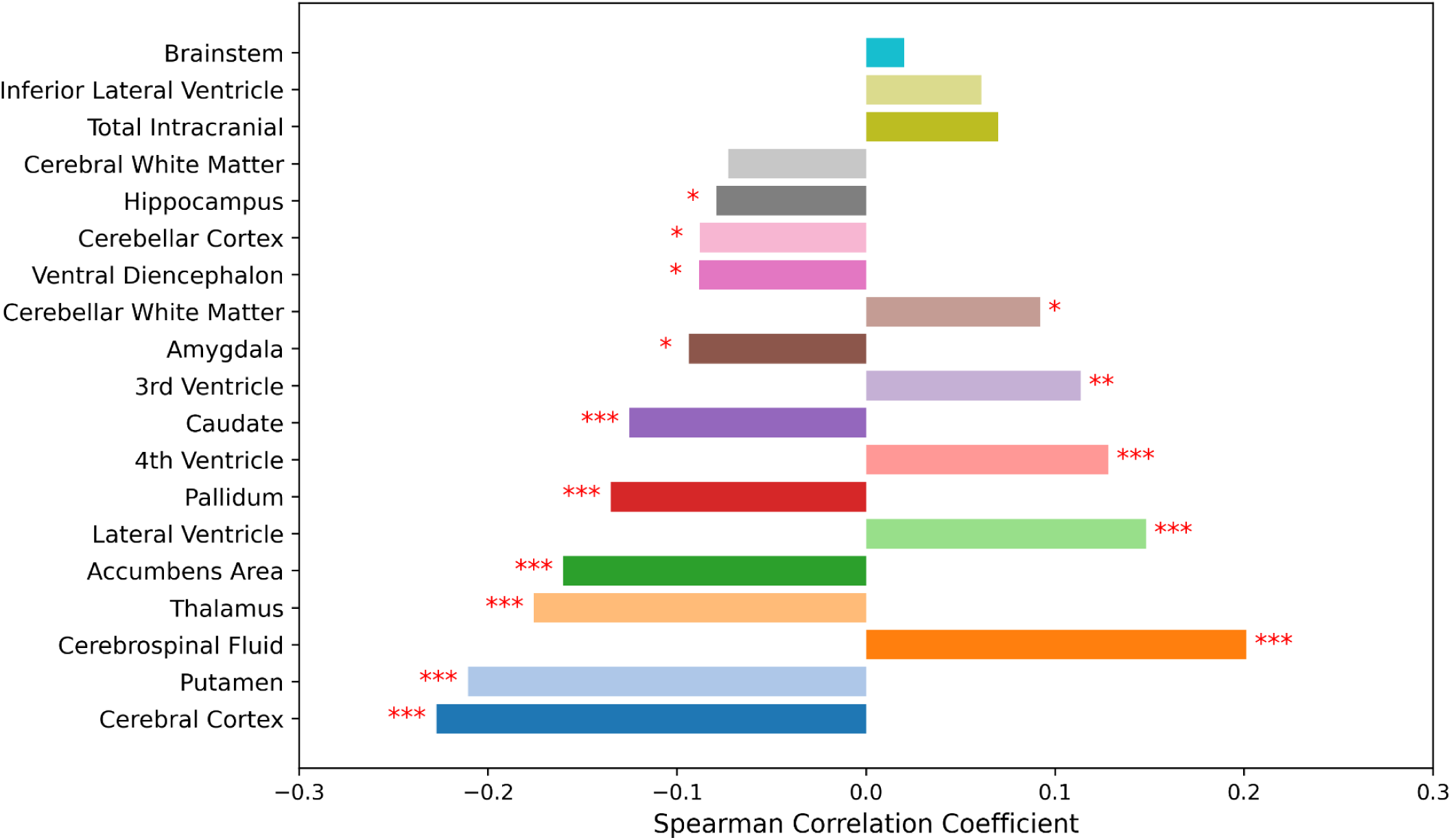
BrainAgeNeXt predictions are strongly correlated with gray matter and cerebrospinal fluid volumes. Correlations between brain parcels (normalized for intracranial brain volume) and BAD. Brain parcellation was performed with SynthSeg (FreeSurfer)^56^. Significance level: * *p* < 0.05, ** *p* < 0.01, *** *p* < 0.001.

### BrainAgeNeXt brain age predictions in people with MS (pwMS)

We predicted BA on our three longitudinal cohorts of pwMS using our proposed BrainAgeNeXt, as it outperformed all other approaches on the HV testing dataset.

#### Cross-sectional analysis

We considered all pwMS from the LCL (3T MRI scans), RADIEMS, and Long 7T cohorts. pwMS had a higher median BAD of 4.21 years (range: −7 to 29, IQR: 8.23) compared to HV in the testing dataset, who had a mean BAD of 0.02 years (range: −13 to 17, IQR: 4.41), *p* < 0.001 (Figure 5). No significant difference was observed in predicted BA between males and females (*p* = 0.09). However, the trend indicated slightly worse performance for men.

**Figure 5.**
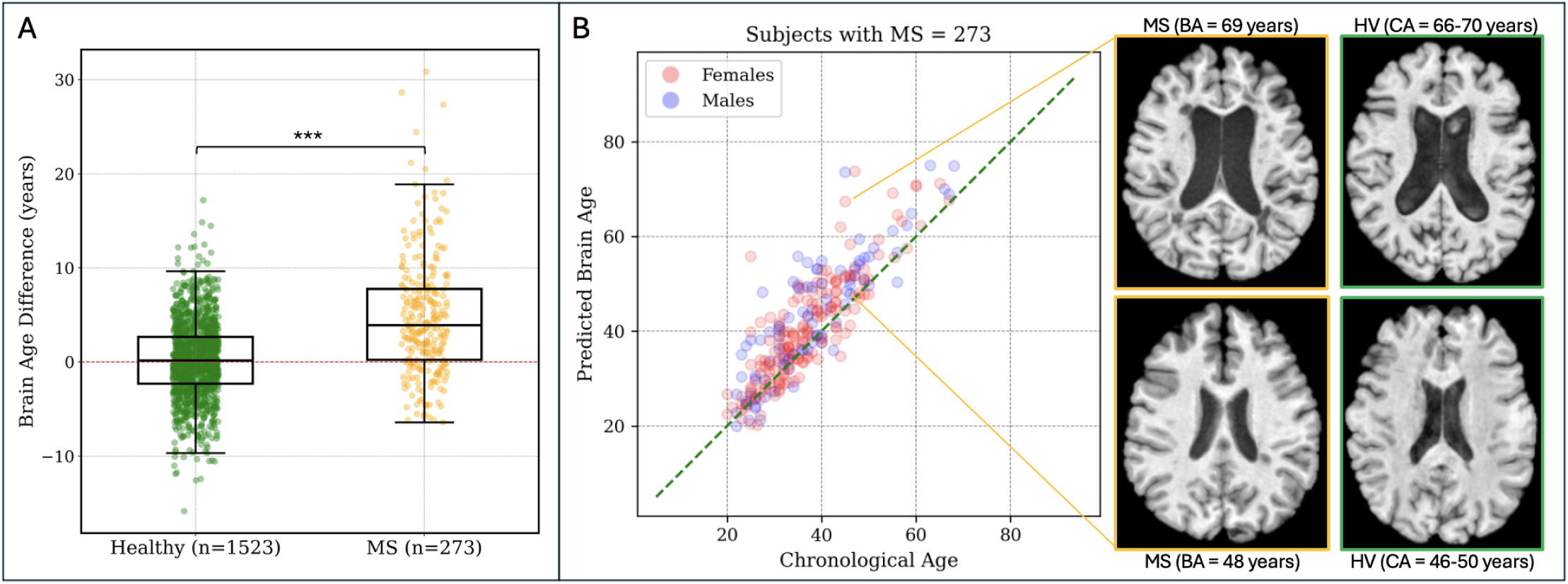
Brain age is greater in individuals with MS compared to HV. A: The predicted brain age (BA) for pwMS is on average 4.20 years greater than their chronological age (CA). Significance level: *** p < 0.001. B: Correlation between predicted brain age and chronological age in individuals with MS. A representative axial MRI slice of two individuals with MS (age range 46-50 years) is shown on the right. The individual with a brain age of 69 years has a greater degree of brain parenchymal atrophy and ventricular enlargement.

We explored the effect of MS lesions on the predicted BA. For a subset of pwMS from the RADIEMS cohort (*n* = 162), we tested BrainAgeNeXt on lesion-filled images (Figure 6A). When comparing the original BA with the BA obtained from lesion-filled images, we observed a strong correlation between the two values (Spearman’s ρ = 0.99). The mean difference between them was minimal (at 0.26 ± 1.32 years), and this difference was not significantly associated with the lesion volume (*p* = 0.08). Thus, we used the non-lesion-filled images to predict BA for the subsequent analyses.

**Figure 6.**
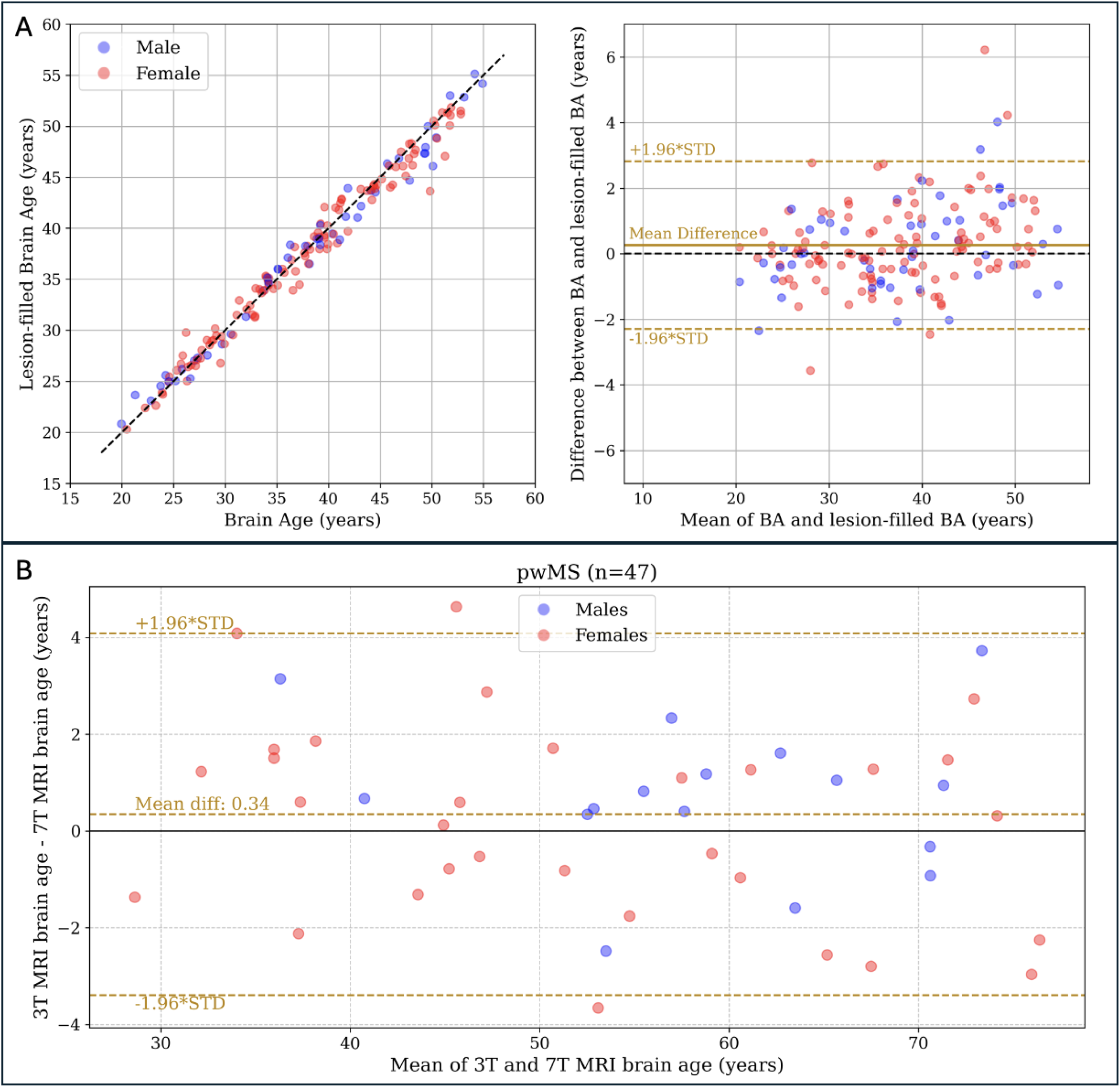
Predicted brain age is similar in lesion filled and non-lesion filled images. A: comparison of the predicted brain age from the original T1w MRI and after performing lesion filling in people with multiple sclerosis (pwMS) from the RADIEMS cohort (*n* = 162). The two values are strongly correlated (Spearman’s ρ = 0.99, *p* < 0.001), with a mean difference of 0.26 years (middle plot, solid golden line). B: Bland-Altman plot illustrating a strong correlation between brain age predictions from 3T and 7T MRI for pwMS in the LCL cohort (n = 47), with Spearman’s ρ = 0.99, p < 0.001, and a mean absolute error (MAE) of 1.5 years.

For the LCL cohort, we predicted BA on both 3T and 7T MRI scans of the same individuals (Figure 6B). The two predictions were strongly correlated (Spearman’s ρ = 0.99, *p* < 0.001) with a mean absolute difference of 1.5 ± 1.0 years and a BAD of 0.3 years (Figure 6B).

We further investigated the correlations between CA, BA, and BAD with MRI measures (Figure 7). Pooling LCL and RADIEMS cohorts together, normalized brain volume (NBV) held a stronger correlation with brain age (r = −0.75, p < 0.001) than chronological age (r = −0.59, p < 0.001). Brain age accounts for 21.8% more variance in NBV compared to chronological age, explaining 56.4% of the variance (95% CI: 47.2% – 64.2%) versus 34.6% (95% CI: 24.3% – 44.4%). In both LCL and RADIEMS cohorts, BA and BAD were negatively associated with NBV, deep gray matter volume, cortical volume, and white matter volume. Both BA and BAD were strongly correlated with white matter lesion volume.

**Figure 7.**
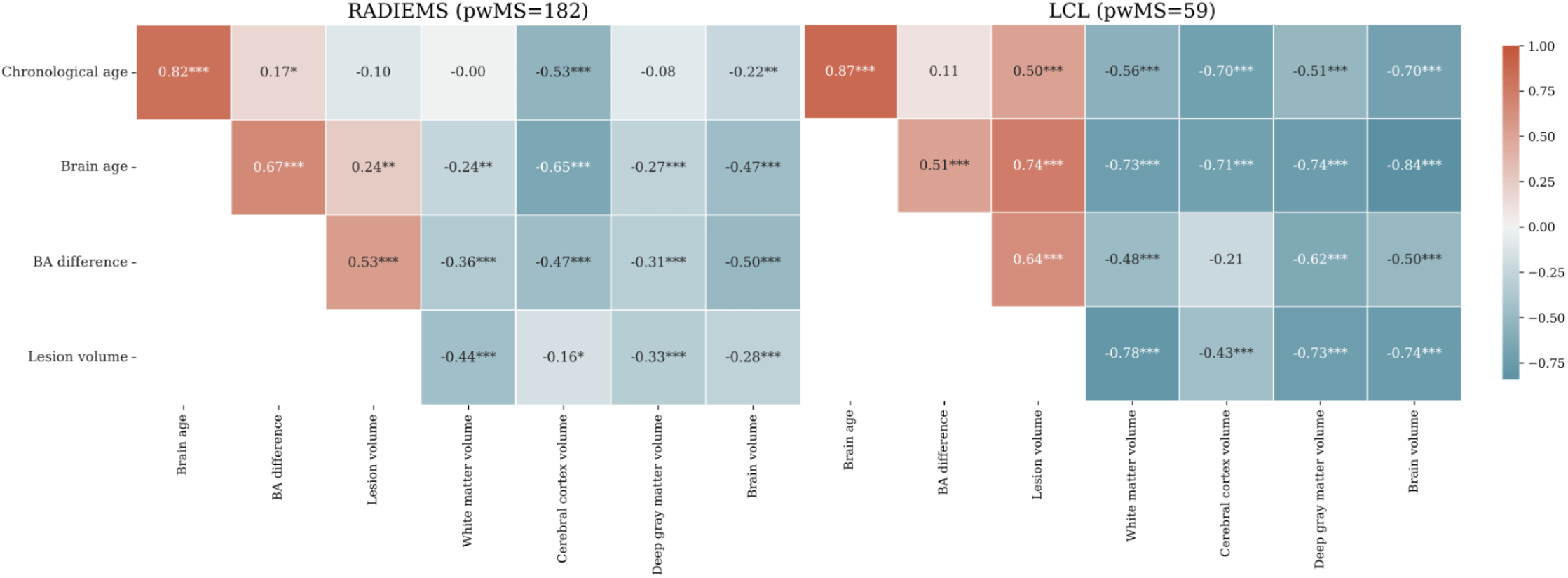
Brain age difference is correlated with brain volumetric measures in pwMS. Heatmap showing Spearman’s correlation coefficient between chronological age, brain age, brain age difference, and MRI markers for the RADIEMS (left) and LCL (right) cohorts. Significance level: * *p* < 0.05, ** *p* < 0.01, *** *p* < 0.001, all adjusted for multiple comparisons using the false discovery rate method.

We then examined the relationship between BAD and its interaction with disease duration on NBV using a generalized linear model, adjusting for dataset effects. The negative association of BAD with NBV (unstandardized B = −0.002 (95% CI: −0.003 to −0.002), p < 0.001) was stronger with longer disease duration (unstandardized B = −5.66 × 10^-5 (95% CI: −9.79 × 10^-5 to −1.53 × 10^-5), *p* = 0.007).

Further, we analyzed relationships between brain age and four measures of disability: EDSS, SDMT, 9HPT, and T25FW (Figure 8). The two MS cohorts exhibited similar trends, but the strength of the associations varied between them, with LCL showing stronger correlations. BAD was significantly correlated with SDMT, EDSS, and 9HPT in both cohorts.

**Figure 8.**
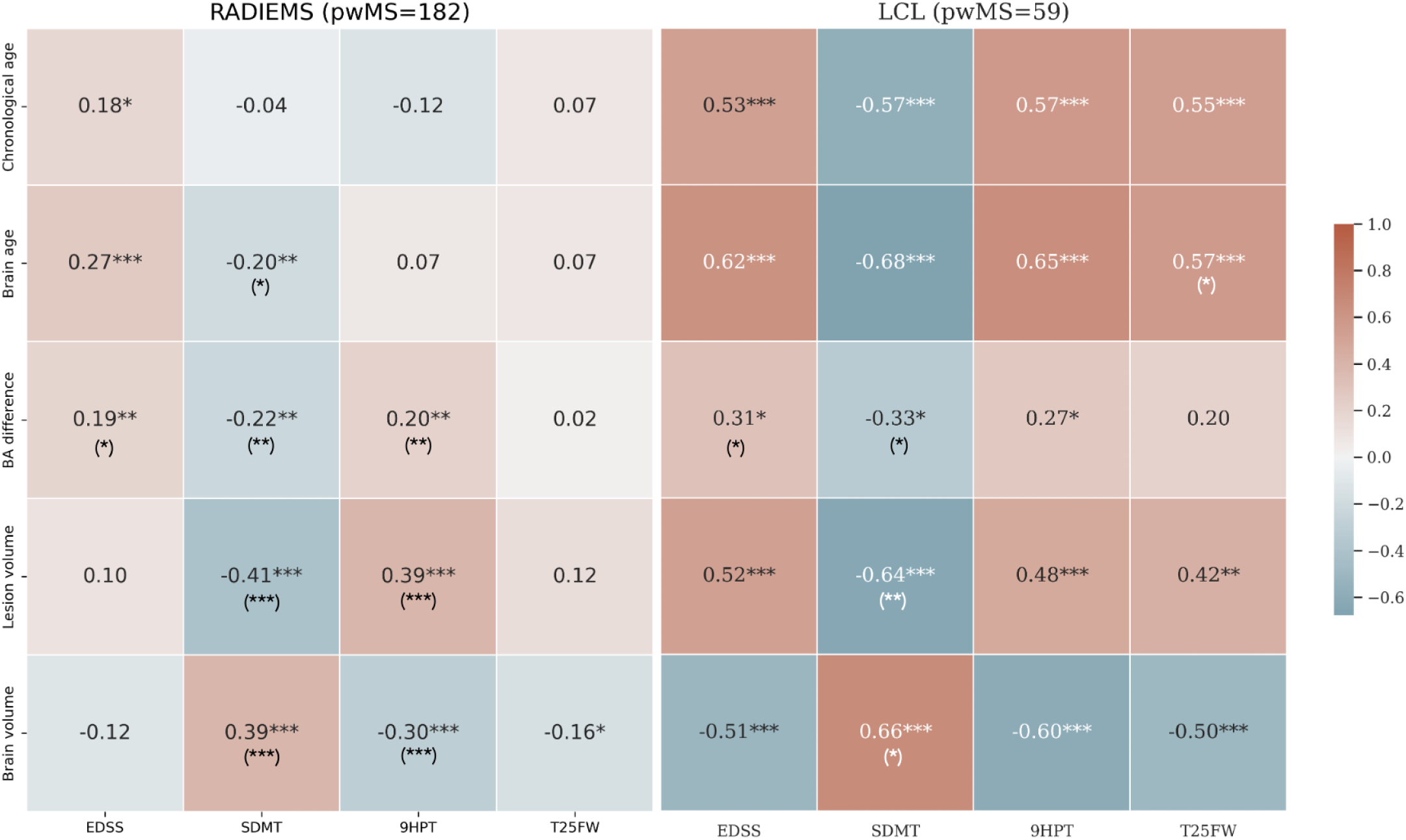
Brain age difference is associated with disability in pwMS. Heatmap showing Spearman’s correlation coefficient between chronological age, brain age, brain age difference, and clinical measures for the LCL and RADIEMS cohorts. EDSS: Expanded Disability Status Scale; SDMT: Symbol Digit Modalities Test; 9HPT: Nine-Hole Peg Test; T25FW: Timed 25-Foot Walk. Significance level: * *p* < 0.05, ** *p* < 0.01, *** *p* < 0.001, all adjusted for multiple comparisons using the false discovery rate method. Asterisks in parentheses indicate significance after adjusting for age.

### Longitudinal analysis

We analyzed the predicted BA for all subjects with MS with at least two sessions (*n* = 200, mean time interval between sessions = 2.85 years, range 1.43−5.51 years). To assess the variability in BA longitudinal change across individual subjects, we computed the annual BA change for each subject separately. The mean annual BA change rate was 1.15 years (IQR 0.96, 1st quartile 0.68, 3rd quartile 1.64), with a 95% confidence interval of 1.05 to 1.26 years. This is statistically different from 1 (one-sample t-test, *p* = 0.003), revealing that in pwMS, brain aging is about 15% faster than chronological aging. The values for each MS cohort are reported in the Supplementary Material (Supplementary Table 1). For 38 subjects from the LCL cohort, both 3T and 7T scans at baseline and follow-up visits were acquired. The subject-wise mean difference between 3T and 7T annual BA change was −0.1 years (Supplementary Figure 1). No significant difference in annual BA change was found between 3T and 7T MRI (*p* = 0.6).

When dividing the pwMS into early MS (disease duration of less than 6 years) and longstanding MS (disease duration greater than 6 years), we observed a statistically significant difference (*p* = 0.004) in the annual brain age change between these two groups (Figure 9A). The early MS group had a mean annual brain age change of 1.24 ± 0.64, compared to 0.75 ± 1.08 for the longstanding MS group.

**Figure 9.**
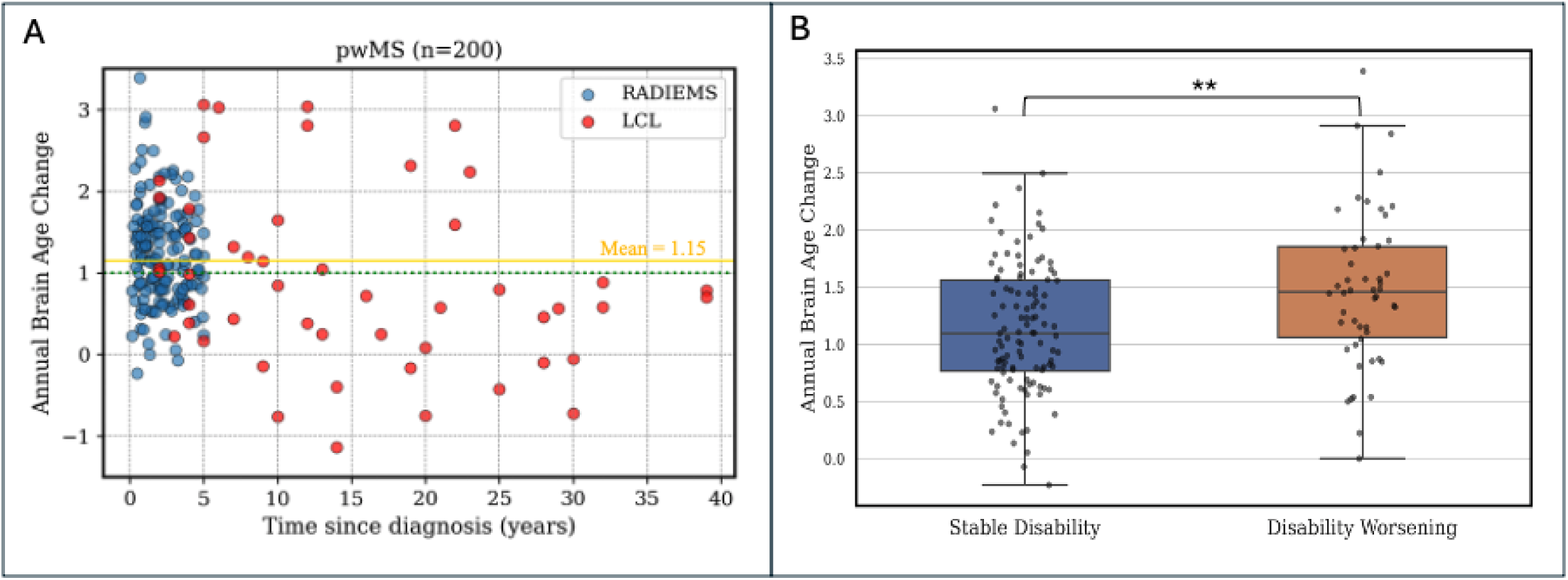
Brain aging is accelerated in MS and is associated with disability worsening. A: the longitudinal brain age analysis reveals that individuals with MS (n=200) have accelerated brain aging (mean annual BA change = 1.15). The green dotted line shows the annual chronological age change, the solid gold line shows the mean annual BA change in pwMS. B: Subjects with early MS (diagnosis <6 years, *n* = 163) are divided into two groups: those showing disability worsening, indicated by an increase in EDSS of ≥1.0 for baseline EDSS <6.0 or ≥0.5 for baseline EDSS ≥6.0, or 20% increase in T25FW, or 20% increase in 9HPT; and those whose clinical assessment remained stable. The subjects who experienced disability worsening between the two visits had a significantly greater brain aging slope (1.24) than those who had no disability worsening (0.75, p < 0.01). Significance level: ** *p* < 0.01.

Additionally, we categorized all pwMS based on disability progression: those exhibiting disability worsening, defined by an EDSS increase of ≥1.0 for baseline EDSS <6.0 or ≥0.5 for baseline EDSS ≥6.0, or a 20% increase in T25FW or 9HPT; and those with stable clinical assessments. In early MS, individuals with disability worsening had a greater BA increase per year (1.48 ± 0.68 years) compared to individuals that remained stable (1.14 ± 0.60 years, *p* < 0.01) (Figure 9b). No difference between the groups was observed in longstanding MS (*p* = 0.08).

## Discussion

In this work, we propose a novel brain age estimation method, coined BrainAgeNeXt, inspired by the state-of-the-art convolutional neural network framework MedNeXt^58,59^. The proposed BrainAgeNeXt model demonstrated superior performance compared to other state-of-the-art approaches in predicting brain age across a large, diverse dataset of over 1’300 HV MRI scans, achieving a competitive MAE of 2.78 years. This highlights the robustness and generalizability of BrainAgeNeXt, even when tested on MRI scans from various sites while preserving individuals’ differences. The findings of our study provide new insights into the application of deep learning-based models for BA estimation, particularly in individuals with MS.

One of the key advantages of BrainAgeNeXt is its ability to learn directly from raw MRI data without relying on pre-extracted volumetric features. This approach mitigates the issues associated with brain parcellation variability and facilitates the model’s applicability to real-world clinical settings. The data augmentation techniques employed during training, including spatial transformations and the simulation of MRI artifacts, further contributed to the model’s robustness, enabling it to generalize well across different levels of image quality. BrainAgeNeXt is the best-performing approach even in the presence of motion artifacts, which considerably degrade the performance of voxel-based morphometry methods such as brainageR^70^. The worse performance of brainageR with motion-degraded images is likely due to the influence of motion on brain parcellation performed by SPM12^67^, which uses a Bayesian framework for tissue classification and anatomical parcellation. Motion artifacts can significantly degrade tissue contrast, making accurate segmentation challenging and brain age estimation less reliable.

Our study also highlights the importance of high-resolution MRI in brain age modeling. To the best of our knowledge, BrainAgeNeXt is the first deep learning-based approach trained with 7T data, resulting in improved performance on 7T brain age predictions compared to other state-of-the-art models. However, whether brain age predictions from 7T MRI are more sensitive to accelerated aging still needs to be explored. 7T MRI also allows the assessment of advanced MS biomarkers, such as cortical lesions, which are very difficult to quantify at lower magnetic field strengths^72^. Thus, 7T MRI-derived brain age could be combined with other MS markers in the future to increase its prognostic value.

We found that brain age difference was correlated with volumes of several key brain structures. Specifically, brain age difference was positively correlated with CSF volume and negatively correlated with cerebral cortex and deep gray matter volumes. This aligns with the known patterns of normal aging process, where increased CSF volume reflects brain atrophy, including shrinking of the cerebral cortex and loss of deep gray matter. These structural changes are indicative of age-related degeneration, suggesting that HV with higher brain age difference values exhibit more pronounced aging-related brain changes. Thus, the ability of BrainAgeNeXt to capture these associations highlights its potential utility in identifying deviations from typical healthy aging trajectories.

Recently, there has been growing interest in brain age in the context of MS^16,17,73^. Several studies have shown that brain age difference is greater in pwMS compared to HV. Brain age difference has also been associated with brain atrophy, physical disability, and cognitive performance, as measured by the SDMT, in pwMS^17^. In two independent longitudinal studies, Cole et al. and Brier et al. found that a higher brain age difference at baseline was associated with greater disability accumulation after a few years^16,73^. This evidence suggests that brain age is a potentially useful metric to measure structural changes in the brain that are associated with disability in MS.

To characterize brain age in MS, we tested BrainAgeNeXt on three longitudinal cohorts of subjects with MS, for a total of 273 individuals. The cross-sectional analysis revealed a significant difference in brain age difference in MS compared to healthy volunteers. Individuals with MS exhibited at baseline an average brain age approximately 4 years greater than their chronological age. Our results are consistent with previous studies^16,17,73^, highlighting the accelerated brain aging process associated with MS. We also analyzed the impact of lesion-filled images on brain age predictions. Although white matter lesion volume was strongly correlated with brain age difference, we found that lesion-filling did not significantly change brain age predictions. This suggests that while MS lesion burden is associated with accelerated aging, the lesions themselves are not the primary drivers of BrainAgeNeXt’s predictions.

Our analysis revealed that brain age is a stronger predictor of normalized brain volume than chronological age across the LCL and RADIEMS cohorts. This stronger correlation suggests that brain age provides a better representation of the neurobiological aging process than chronological age in pwMS. This finding highlights the added value of brain age over chronological age in assessing pwMS, as it reflects the underlying pathological processes more accurately. Additionally, both brain age and brain age difference are negatively associated with the normalized volume of key brain structures, including deep gray matter, cortex, and white matter. This consistent pattern of atrophy highlights the combined impact of aging and neurodegeneration in MS.

Importantly, the interaction between BAD and disease duration reveals a compounded negative effect on normalized brain volume. As disease duration increases, the association between brain age difference and brain volume becomes more pronounced. This suggests that subjects who have had MS for a longer time experience a greater degree of brain atrophy. Our results, therefore, emphasize the progressive nature of brain atrophy in MS, influenced not only by the accelerated brain aging process but also by the disease duration.

The longitudinal brain age analysis demonstrates that, on average, brain age tends to increase at a considerably faster rate than chronological age in early MS (<6 years since diagnosis), while this annual change may decrease in the later stages of the disease. Larger studies, however, are needed to confirm these initial findings. Among all subjects with MS, the annual brain age change showed a wide IQR of 0.96. This highlights an inter-subject variability in brain aging rates, the drivers of which will be important to explore in the future. This provides additional evidence for the potential utility of brain age modeling in monitoring disease progression, particularly at its early stages. No significant differences were found when comparing the annual change in brain age predicted from 3T and 7T MRI scans. This confirms the robustness of BrainAgeNeXt across different magnetic field strengths.

The brain age difference is associated with several disability measures in both early and longstanding MS, suggesting that brain age may offer valuable prognostic information. The strongest correlations were found with SDMT, in line with previous studies^16,17^. Notably, brain age was more closely associated with all disability measures than chronological age. Additionally, in early MS, a greater annual brain age increase was linked to worsening disability. In the future, by identifying individuals with accelerated brain aging, clinicians may be able to implement early interventions and personalized treatment plans aimed at mitigating the progression of cognitive impairment. Moreover, longitudinal brain age estimation could serve as an endpoint in clinical trials to assess the efficacy of therapeutic interventions and lifestyle modifications.

Despite the promising results, several limitations should be acknowledged. The healthy volunteers and MS datasets used for training and validation predominantly consisted of European and American individuals, which may limit the generalizability of our findings to other ethnicities and populations. Future studies should aim to include more diverse cohorts to ensure the broad applicability of brain age models. Additionally, while BrainAgeNeXt demonstrated excellent performance in predicting brain age, the clinical utility of brain age as a prognostic biomarker for MS requires further validation through longitudinal studies with larger cohorts and testing in the context of interventional clinical trials. Finally, the performance of BrainAgeNeXt should be evaluated in future studies using clinical-grade scans and other T1w sequences.

In conclusion, BrainAgeNeXt, which we have made publicly available^2^, represents a significant advance in brain age modeling. It offers a robust and accurate method for estimating brain age from diverse MRI data, including 7T MRI. Its application to individuals with MS demonstrates the potential for brain age as a valuable biomarker for assessing accelerated aging and MS disability progression. By requiring only a T1-weighted MRI to compute brain age, we hope to advance brain age research further and facilitate its application to neurological conditions.

## Data Availability

All data produced in the present study are available upon reasonable request to the authors.

https://github.com/FrancescoLR/BrainAgeNeXt

## Footnotes

1 https://github.com/FrancescoLR/BrainAgeNeXt

2 https://github.com/FrancescoLR/BrainAgeNeXt

