## Supplementary for "BrainAgeNeXt: Advancing Brain Age Modeling for Individuals with Multiple Sclerosis"

### Supplementary Material

| MS Cohort | Time since diagnosis (years) | Time between MRI sessions (years) | BA Change per year (years) |
|---|---|---|---|
| RADIEMS | 2.23 ± 1.46 | 3.21 ± 0.40 | 1.23 ± 0.62 |
| LCL | 14.22 ± 10.86 | 2.18 ± 0.51 | 0.90 ± 1.06 |

**Supplementary Table 1. Brain age increases more rapidly at the early stages of multiple sclerosis.**

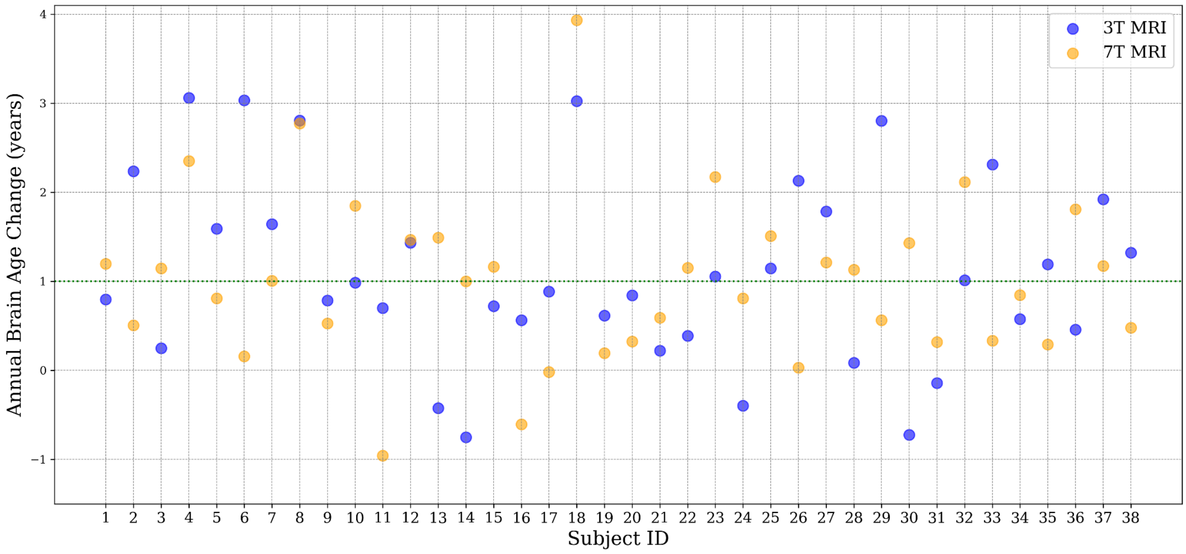


**Supplementary Figure 1. The annual change in brain age does not significantly differ when comparing 3T and 7T MRI brain age (*p*=0.6).** The plot shows a comparison of 38 pwMS from the LCL cohort imaged at both 3T and 7T.

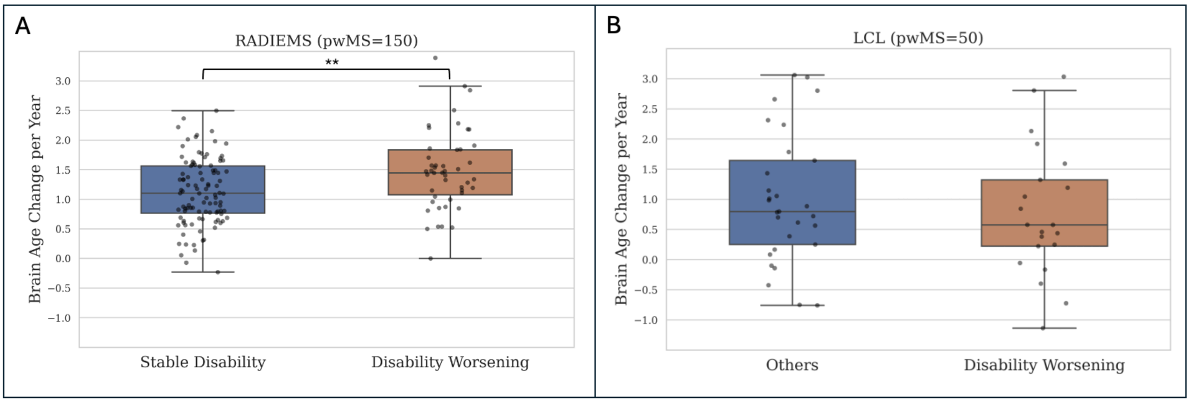


**Supplementary Figure 2. The annual brain age change is greater for subjects with disability worsening compared to stable ones in RADIEMS, whereas no differences are observed in LCL.** Significance level: ** $p < 0.01$.
